# Using fluorescently labeled vedolizumab to visualize local drug distribution during colonoscopy and identify mucosal target cells in patients with inflammatory bowel disease

**DOI:** 10.1101/2023.10.25.23297524

**Authors:** Ruben Y. Gabriëls, Anne M. van der Waaij, Matthijs D. Linssen, Michael Dobosz, Pia Volkmer, Sumreen Jalal, Dominic J. Robinson, Marcela A. Hermoso, Marjolijn N. Lub-de Hooge, Eleonora A.M. Festen, Gursah Kats-Ugurlu, Gerard Dijkstra, Wouter B. Nagengast

## Abstract

**Background:** Improving patient selection and development of biological therapies such as vedolizumab in inflammatory bowel disease (IBD) requires a thorough understanding of the mechanism of action and target binding, thereby providing individualized treatment strategies. Our goal was to visualize the macroscopic and microscopic distribution of intravenous injected fluorescently labeled vedolizumab, vedo-800CW, and identify its target cells using fluorescence molecular imaging (FMI).

**Methods:** In total 43 FMI procedures were performed in 37 IBD patients. FMI procedures consisted of macroscopic in vivo assessment during endoscopy, followed by macroscopic and microscopic ex vivo imaging. In phase A patients received a dose of 4·5 mg or 15 mg vedo-800CW or no tracer prior to endoscopy. In phase B patients received 15 mg vedo-800CW preceded by an unlabelled (sub)therapeutic dose of vedolizumab.

**Findings:** FMI quantification showed a significant dose-dependent increase in vedo-800CW fluorescence intensity in inflamed tissues, with 15 mg (153·7 a.u. [132·3-163·7]) as most suitable tracer dose compared to 4·5 mg (55·3 a.u. [33·6-78·2]) in naïve patients (p=0·0002). Moreover, the fluorescence signal decreased by 61% when vedo-800CW was administered after a therapeutic dose of unlabeled vedolizumab, suggesting target saturation in the inflamed tissue. Fluorescence microscopy and immunostaining showed that vedolizumab penetrated the inflamed mucosa and was associated with several immune cell types. Finally, surface binding of vedo-800CW was most prominent in plasma cells, whereas intracellular localization was observed primarily in macrophages and eosinophils.

**Interpretation:** These results indicate the potential of FMI to macroscopically determine the local distribution of drugs in the inflamed target tissue and identify drug target cells, providing new insights into targeted agents for their use in IBD. Regarding vedolizumab, we provide valuable information about its main target cells, contributing to our understanding of the underlying mechanism of action.

**Funding:** This work received funding from the EU/EFPIA/IMI2 JU Immune-Image grant no831514.

**Research in context:** *Evidence before this study:* Combining fluorescence molecular imaging (FMI) with fluorescently labeled drugs holds high potential for providing detailed insights into the drug’s mechanism of action by allowing researchers to visualize its distribution and its target cells. Strikingly, with respect to inflammatory bowel disease (IBD) our lack of understanding regarding the mechanism of action of therapeutic compounds such as vedolizumab remains a major hurdle to improving prognosis and quality of life. Vedolizumab inhibits α4β7 integrin and was developed to prevent the migration of α4β7-expressing gut-homing T cells from vessels into the mucosa, thereby preventing inflammation. However, recent studies have speculated that the anti-inflammatory effect of vedolizumab is mediated by a wide range of α4β7-expressing immune cells, not just T cells. Unfortunately, a literature search revealed that drug distribution studies on vedolizumab in IBD that examined the mucosal distribution of vedolizumab or its target cells are lacking.

*Added value of this study:* Here, we show for the first time that intravenous administration of a fluorescently labelled drug can be used to visualize both the macroscopic and microscopic tissue distribution using FMI. Importantly, we combined fluorescently labeled vedolizumab with FMI in 43 procedures in patients with IBD, revealing valuable information regarding the drug’s distribution. We performed both *in vivo* and *ex vivo* FMI in order to quantify vedolizumab levels in inflamed mucosal tissues and found that vedolizumab targets a variety of immune cell types. We examined subcellular localization in these immune cells in more detail and found that vedolizumab binds to the surface of plasma cells, but is taken up into the cytoplasm in macrophages and eosinophils. These findings provide proof-of-concept to support the notion that FMI can be used to determine the distribution of a drug in the target tissue and identify the drug’s cellular target. Using this novel imaging technique will additionally provide valuable new insights regarding a drug’s ideal dose and the target saturation of specific drugs used to treat inflammatory disease.

*Implications of all the available evidence:* The ability to localize a drug’s distribution and identify its target cells is an essential step towards improving treatment options for IBD and other inflammatory diseases, thereby eventually improving outcome and increasing quality of life. Our step-by-step FMI approach consisting of *in vivo* macroscopic fluorescence imaging, *ex vivo* fluorescence tissue analysis, and fluorescence microscopy can be used to increase our understanding of drug distribution at the target levels and thereby help understanding the underlying mechanism of action for a wide range of drugs. Ultimately, these findings may help minimize the economic and social impacts of chronic inflammatory diseases.

## Introduction

Biological therapies such as the monoclonal antibody vedolizumab are important options for the treatment of inflammatory bowel disease (IBD)^1^, a group of chronic idiopathic inflammatory disorders that affect the gastrointestinal tract and consist primarily of Crohn’s disease (CD) and ulcerative colitis (UC). Unfortunately, only half of all patients with IBD respond to vedolizumab therapy, and only 39-45% of patients maintain clinical remission.^2^ Selecting patients who are suitable for vedolizumab therapy is currently not possible, as no reliable tools are available for predicting response. Moreover, administering an unsuitable treatment can lead to unnecessary disease burden, adverse effects, and high costs^3^, thus highlighting the need to develop clinical tools for optimized treatment and to predict response in individual patients. Several parameters have been investigated as potential indicators of patient outcome, including serological biomarkers, mucosal biomarkers, vedolizumab trough levels, clinical scores, and the gut microbiome; however, none has shown a sufficient correlation with the response to vedolizumab therapy in order to be incorporated into the clinical decision-making process.^4–7^ In addition, surprisingly little is known regarding drug distribution in the inflamed gut.

Currently, vedolizumab’s precise mechanism of action is under debate. Initially, the prevailing theory was that vedolizumab reduces inflammation by specifically binding to the α4β7 integrin expressed on gut-homing T cells. The resulting integrin-antibody complex was believed to block T cells from trafficking to the gastrointestinal mucosa, thereby preventing inflammation.^8–10^ In contrast, recent studies suggest that a wide range of α4β7-expressing immune cells may be involved in mediating vedolizumab’s anti-inflammatory effects.^11–19^ However, none of these experimental studies directly visualized the tissue distribution of vedolizumab or its interaction with target cells in the inflamed gut mucosa.

Previously, we used fluorescence molecular imaging (FMI) to visualize the distribution of fluorescent tracers in gastrointestinal malignancies.^20,21^ FMI is a relatively new imaging technique that includes both *in vivo* fluorescence imaging during endoscopy and *ex vivo* fluorescence analysis of tissue samples; however, to date this technique has not been used to study drug distribution in inflammatory disease. Here, we performed a phase 1 clinical trial to assess the feasibility of using FMI after intravenous administration of fluorescently labeled vedolizumab in order to visualize the drug’s distribution and identify potential target cells in patients with IBD.

## Materials and Methods

### Study design

This single-center phase 1 clinical feasibility trial was performed at the University Medical Center Groningen (UMCG). The study was approved by the UMCG’s Institutional Review Board (METc Groningen; 2019/305) and was conducted in accordance with the Dutch Act on Medical Research involving Human Subjects (WMO) and the principles of the Declaration of Helsinki (adapted at the 64th WMA General Assembly in Fortaleza, Brazil, 2013). The trial was registered at ClinicalTrials.gov (NCT04112212).

Both vedolizumab-naïve patients with IBD and patients with IBD who received vedolizumab therapy for at least 14 weeks were eligible to participate in the study. Written informed consent was obtained from all patients. To meet the inclusion criteria, patients had to be ≥18 years of age, have an established diagnosis of IBD, and be eligible to receive vedolizumab therapy. We excluded female patients who were pregnant or breastfeeding. All study-related procedures are depicted in Figure 1.

**Figure 1:**
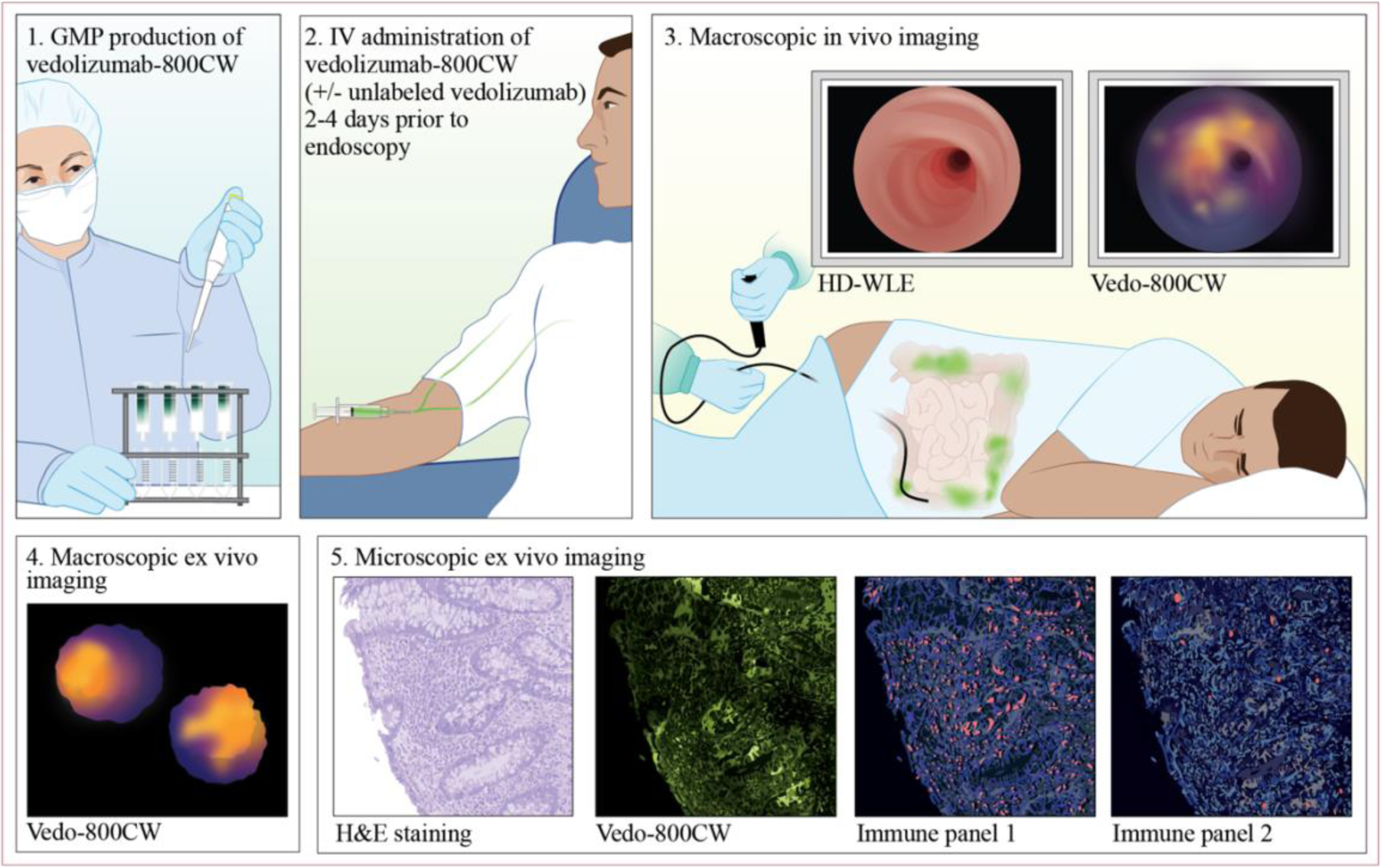
overview of the procedures included in this study. (1) The fluorescent tracer vedo-800CW was produced in accordance with good manufacturing practices (GMP). (2) Vedo-800CW with or without unlabeled vedolizumab (where applicable) was administered intravenously 2-4 days prior to endoscopy. (3) A tandem in vivo procedure including high-definition white-light endoscopy (HD-WLE) and fluorescence molecular endoscopy was performed in order to image the drug’s distribution. (4) Fluorescence intensity was quantified ex vivo on formalin fixed paraffin embedded (FFPE) blocks in all biopsies. (5) Finally, 4 μm tissue sections were used to assess histopathological inflammation status based on H&E staining, to visualize the microscopic drug distribution, and to identify immune target cell types.

### GMP production of vedo-800CW

The fluorescent tracer vedolizumab-800CW (vedo-800CW) was produced by conjugating vedolizumab (Entyvio; Takeda Pharma, Tokyo, Japan) to the IRDye 800CW NHS ester near-infrared dye (LI-COR Biosciences, Lincoln, NE) in the UMCG Department of Clinical Pharmacy and Pharmacology’s GMP production unit in accordance with EU GMP guidelines. A detailed description of the labelling and development process was reported previously.^22^

### Patient cohorts

This clinical trial consisted of five distinct patient cohorts and was divided into two phases (A and B), as shown schematically in Figure 2. In phase A, 15 vedolizumab-naïve patients were enrolled in a vedo-800CW dose-finding study. Each patient was then assigned to one of the following vedo-800CW dose cohorts (*n*=5 patients each): 0 mg (serving as a negative control group), 4·5 mg, or 15 mg vedo-800CW. An interim analysis was conducted in order to evaluate the best dose by assessing various safety parameters and imaging results. Based on this interim analysis, the group that received 15 mg was increased by including 10 additional patients, for a total of 15 patients in this group. In phase B, a dose of 15 mg was then administered to two additional patient cohorts in order to evaluate mucosal saturation of the drug. First, 5 vedolizumab-naïve patients received a single subtherapeutic dose (75 mg) of unlabeled vedolizumab followed by a 15 mg dose of vedo-800CW. Next, 13 patients received a 15 mg dose of vedo-800CW after a routine vedolizumab treatment infusion; 12 of these 13 patients received 300 mg vedolizumab in accordance with current guidelines, while the remaining patient received a higher dose of 600 mg due to their BMI of 61·8. Three patients in the therapeutic dosing group received the tracer after receiving their first dose in the therapeutic vedolizumab regimen; the remaining ten patients had received at least 14 weeks of vedolizumab treatment prior to receiving vedo-800CW. Unlabeled vedolizumab was administered one hour prior to vedo-800CW and both were administered intravenously 2-4 days prior to endoscopy.

**Figure 2:**
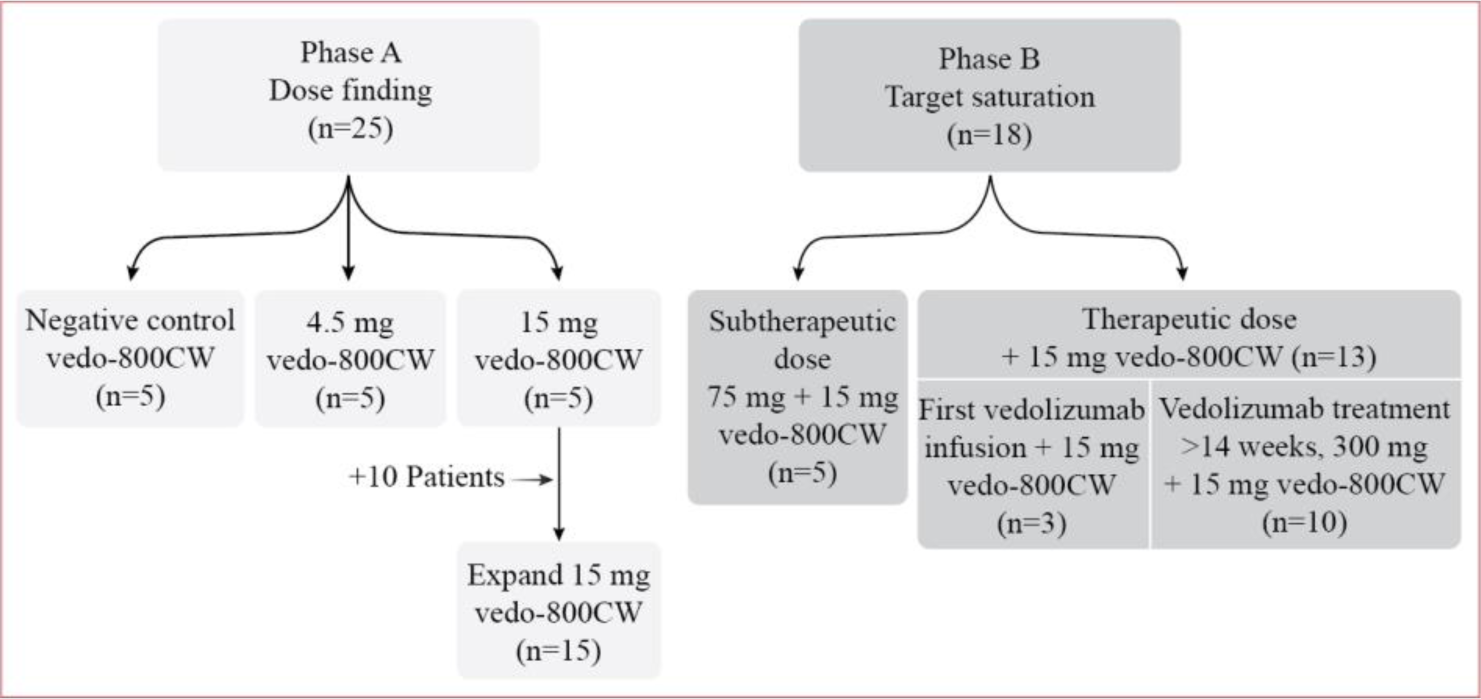
Schematic diagram depicting the various patient cohorts included in phase A and phase B of the study. In total, 43 FMI procedures were performed. In phase A, three doses of vedo-800CW (0, 4·5, and 15 mg) were administered to vedolizumab-naïve patients (n=5 patients each). The 15 mg cohort was then increased to 15 patients. In phase B, both a subtherapeutic dose (75 mg) and a therapeutic dose (300 or 600 mg) of unlabeled vedolizumab was administered, followed by a 15 mg dose of vedo-800CW. Note that 6 patients participated in both phase A and phase B; thus, a total of 37 patients were included in this trial.

Baseline patient characteristics included the Simple Clinical Colitis Activity Index (SCCAI) and the Harvey-Bradshaw Index (HBI) for patients with UC and CD, respectively. In addition, blood and stool samples were collected from all patients and used to perform a complete blood count and to measure plasma C-reactive protein (CRP) and fecal calprotectin levels.

### Macroscopic in vivo imaging

All study-related endoscopy procedures were performed by a gastroenterologist (author WBN) who was experienced in fluorescence endoscopy procedures. A tandem procedure was performed combining high-definition white light endoscopy (HD-WLE) to assess inflammation status with *in vivo* FMI to visualize the fluorescent signals in select ileocolonic segments in endoscopically assessed inflamed and non-inflamed regions. In addition, several biopsies were collected from each section of bowel for subsequent *ex vivo* FMI analyses.

### Macroscopic ex vivo imaging and quantification

All formalin-fixed, paraffin-embedded (FFPE) biopsies were scanned using an Odyssey CLx flatbed scanner (LI-COR Biosciences) to generate fluorescence images. These images were then analyzed by drawing a region of interest around the complete biopsy using the ImageJ package Fiji^23^, which was used to calculate mean fluorescence intensity (FI_mean_). All biopsies were then cut into 4 μm sections for histopathological examination. Specifically, the sections were stained with hematoxylin and eosin (H&E) and used to assess histopathological inflammation severity by an expert gastrointestinal pathologist who was blinded with respect to the patient cohort. The endoscopic assessment of inflammation status was correlated to FI_mean_.

### Microscopic ex vivo imaging

Fluorescence microscopy was used to study the tissue penetration, distribution, accumulation, and potential drug-target interactions of vedo-800CW at cellular resolution. For drug and immunofluorescence imaging, select 4 μm FFPE tissue sections were deparaffinized, mounted on glass slides using Prolong Gold Antifade with DAPI mounting medium (Thermo Fisher Scientific), coverslipped, and scanned using a Zeiss AxioScan Z1 slide scanner. To investigate the potential interaction between vedo-800CW and specific cell types in the innate and adaptive immune system, the coverslips on previously scanned tissue sections were removed by soaking the slides for 30 min in warm water, and two different fluorescent multiplex immunohistochemistry stainings (immune panel 1 and immune panel 2) were performed on serial sections using the Ventana Discovery Ultra Platform (Roche Tissue Diagnostics). A wide range of immune cell types, including adaptive (CD3+ T cells, CD8+ T cells, regulatory T cells, B cells, and plasma cells) and innate (dendritic cells, macrophages, eosinophils, and neutrophils) immune cells, were chosen as the target cells. Finally, the digital images were analyzed using the HALO image analysis platform (Indica Labs, Albuquerque, NM) in order to visualize the tissue distribution of vedo-800CW and to investigate drug-target interactions in specific immune cell subsets. More detailed information on the markers, antibodies used for immunofluorescence, and aimed target cells is presented in Table S1.

### Integrity of the vedo-800CW tracer

SDS-PAGE was performed on fresh-frozen patient biopsy samples in order to determine the stability and integrity of the vedo-800CW conjugate. In brief, sample lysates were separated on 7·5% Mini-PROTEAN TGX Precast Gels (Bio-Rad) using a running buffer consisting of Trizma base, glycine, and 20% (w/v) SDS at 60 V for 2 h; two Lonza ProSieve Color Protein Markers (Fischer Scientific) ranging from 10-190 kDa (cat. BMA50550) and 4·6-300 kDa (cat. BMA00193837) were in cluded as size markers. The gels were scanned using an Odyssey CLx flatbed scanner (LI-COR Biosciences), and fluorescence was visualized at 800 nm.

### Statistical analysis

Given the relatively small sample size, all data are considered to be non-normally distributed. Except where indicated otherwise, summary data are presented as the median and interquartile range. Differences between two groups were analyzed using the Mann-Whitney *U* test (for independent data) or the Wilcoxon signed rank test (for paired data), and a two-sided *p*-value <0·05 was considered to indicate statistical significance. The data were analyzed and plots were generated using Prism 9 (GraphPad Software Inc., San Diego, CA).

## Results

A total of 38 patients with IBD were initially included between February 2020 and April 2022. One patient was subsequently excluded from the study because tracer administration was terminated halfway through the infusion due to a CTCAE (Common Terminology Criteria for Adverse Events) grade I adverse event (headache); thus, 37 patients were included in the study and final analysis. The patient and disease characteristics of each cohort at baseline are summarized in Table 1. Six of the 37 patients in the study participated in both phase A and phase B; thus, in total 43 FMI procedures were completed.

**Table 1:** Patient characteristics at baseline.

|  | Phase A |  |  | Phase B |  |
| --- | --- | --- | --- | --- | --- |
| Characteristic | 0 mg<br>n=5 | 4.5 mg<br>n=5 | 15 mg<br>n=15 | 75 + 15 mg<br>n=5 | Therapy + 15 mg<br>n=13 |
| Age, years | 45 [24-53] | 42 [30-57] | 31 [27-49] | 43 [41-57] | 46 [37-53] |
| BMI | 26.6 [21.2-28.6] | 24.8 [24.6-27.9] | 23.4 [21.9-26.9] | 29.4 [22.0-33.8] | 25.4 [23.0-28.6] |
| **IBD diagnosis** |  |  |  |  |  |
| CD, n (%) | 1 (20%) | 2 (40%) | 7 (47%) | 4 (80%) | 6 (46%) |
| UC, n (%) | 4 (80%) | 3 (60%) | 8 (53%) | 1 (20%) | 7 (54%) |
| **Disease activity based on HBI (CD cases) or SCCAI (UC cases)** |  |  |  |  |  |
| Remission, n (%) | 1 (20%) | 2 (40%) | 4 (27%) | 2 (40%) | 2 (15%) |
| Active disease, n (%) | 4 (80%) | 3 (60%) | 11 (73%) | 3 (60%) | 11 (85%) |
| **Endoscopic assessment of inflammation status** |  |  |  |  |  |
| Non-inflamed, n (%) | 0 (0%) | 0 (0%) | 2 (13%) | 1 (20%) | 2 (15%) |
| Active inflammation, n (%) | 3 (60%) | 1 (20%) | 3 (20%) | 0 (0%) | 2 (15%) |
| Both non-inflamed and active inflammation, n (%) | 2 (40%) | 4 (80%) | 10 (67%) | 4 (80%) | 9 (69%) |
| **Laboratory parameters** |  |  |  |  |  |
| Hemoglobin (mmol/L) | 8.4 [7.0-8.4] | 8.9 [8.6-9.1] | 8.6 [8.1-8.9] | 8.0 [7.7-8.4] | 7.8 [7.5-8.6] |
| CRP (mg/L) | 7.0 [0.9-16.0] | 2.2 [0.5-3.4] | 2.6 [1.1-6.0] | 16.0 [2.8-16.0] | 1.8 [0.6-7.0] |
| Thrombocytes (x10<sup>9</sup>/L) | 329 [273-388] | 308 [308-407] | 318 [247 – 363] | 297 [286-322] | 278 [246-304] |
| Eosinophils (x10<sup>9</sup>/L) | 0.13 [0.07-0.24] | 0.07 [0.01-0.13] | 0.13 [0.09-0.19] | 0.13 [0.09-0.14] | 0.12 [0.05 -0.23] |
| Fecal calprotectin | 635 [425-3770] | 233 [124-473] | 605 [295-1710] | 160 [100-335] | 215 [41-2320] |

*Except where indicated otherwise, data are presented as the median [interquartile range]*
*BMI: body mass index; CD: Crohn's disease; CRP, C-reactive protein; HBI: Harvey-Bradshaw Index; IBD: inflammatory bowel disease; SCCAI: Simple Clinical Colitis Activity Index; UC: ulcerative colitis.*

### Macroscopic in vivo and ex vivo imaging and quantification of vedo-800CW

In phase A of this clinical trial, a conventional endoscopic assessment of gut inflammation status showed active inflammation in 32 out of 37 patients. Real-time *in vivo* macroscopic fluorescence revealed clear uptake of the fluorescent tracer in the target organ—the inflamed gut—compared to healthy (i.e., non-inflamed) gut mucosa (Figure 3A). Semi-quantitative *ex vivo* FI_mean_ was then used to determine the levels of vedo-800CW fluorescence in both the active inflamed and non-inflamed tissues. FI_mean_ in the active inflamed tissue was significantly higher in the 15 mg dose group in phase A compared to the 4·5 mg dose group, with median [IQR] FI_mean_ values of 153·7 a.u. [132·3-163·7] and 55·3 a.u. [33·6-78·2], respectively (*p=*0·0002) (Figure 3B). Negligible signals were found in the control group. Based on this FI_mean_ analysis, 15 mg was chosen for phase B due to the high contrast between inflamed and non-inflamed tissues compared to the 4·5 mg group; this cohort was then expanded to 15 patients by including an additional 10 patients (see Figure 2). Furthermore, the 15 mg cohort was analyzed based on IBD subtype (UC or CD, with 7 and 8 patients, respectively) as well as ileocolonic location (left-sided, right-sided), revealing no significant difference in FI_mean_ between subgroups (Figure 3C). In addition, an SDS-PAGE analysis of fresh-frozen biopsy samples obtained from the 15 mg vedo-800CW cohort confirmed the tracer’s stability and integrity (Figure S1).

**Figure 3:**
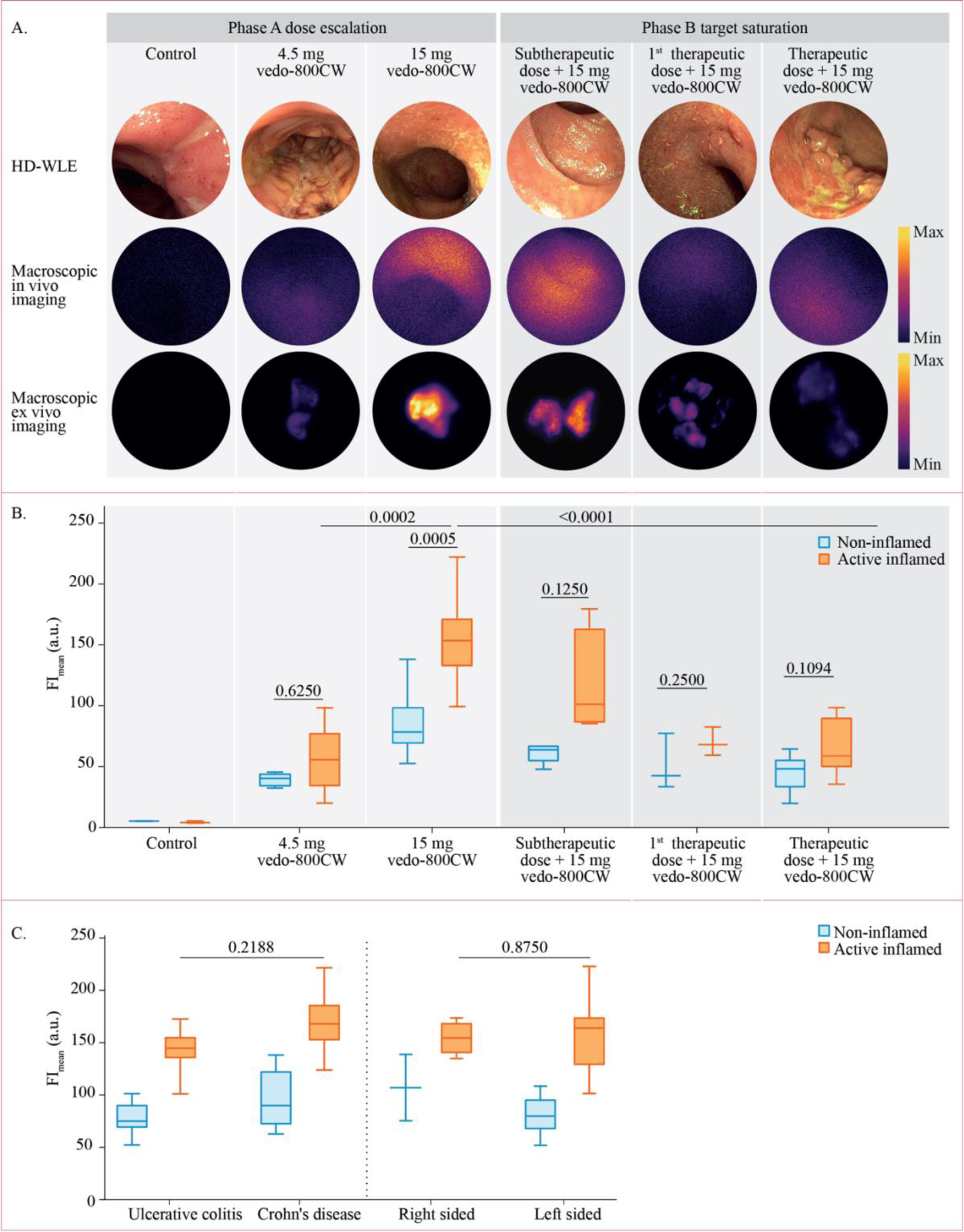
In vivo visualization and ex vivo quantification. (A) Representative high-definition white-light endoscopy (HD-WLE) images (top row) of active inflamed tissue, with corresponding in vivo (middle row) and ex vivo (bottom row) fluorescence images in the indicated cohorts. All fluorescence images were scaled per modality in relation to one another to allow inter-image comparisons. (B) Box plot summarizing FI_mean_ calculated from fluorescence scans of all biopsies in all cohorts, with p-values indicated (Mann-Whitney U test). (C) Box plot summarizing the results obtained from the 15 mg cohort in phase A, stratified by disease subtype and disease location, with p-values indicated (Mann-Whitney U test).

Next, in phase B we assessed the effect of administering unlabeled vedolizumab on the fluorescent signal. Our *ex vivo* FI_mean_ analysis revealed a dose-dependent decrease in signal intensity in active inflamed tissues in patients pretreated with increasing doses of unlabeled vedolizumab followed by vedo-800CW. Specifically, patients who received a subtherapeutic dose (75 mg) followed by 15 mg vedo-800CW had a median [IQR] FI_mean_ of 101·6 a.u. [85·7-163·7]. Patients who received their first therapeutic dose followed by 15 mg vedo-800CW had a FI_mean_ of 69·2 a.u. [60·2-82·4], and patients who received their therapeutic dose after >14 weeks of therapy followed by 15 mg vedo-800CW had a median FI_mean_ of 59·3 a.u. [50·1-90·7]. Compared to the vedolizumab-naïve patients in the 15 mg vedo-800CW cohort in phase A, FI_mean_ was significantly reduced by more than 61% (*p*<0·0001) in the therapeutic dose cohort in phase B (Figure 3A, B). Moreover, FI_mean_ in the therapeutic dose cohort was decreased to the same level measured in non-inflamed tissues in these patients, suggesting that vedo-800CW binding was blocked by unlabeled vedolizumab in the inflamed mucosa.

### Microscopic analysis of drug distribution and drug-target interaction

Next, we performed *ex vivo* fluorescence microscopy in order to assess the distribution of vedo-800CW in both 20 μm, 10 μm and 4 μm tissue sections (Figure S2). Our analysis revealed deep penetration and a heterogeneous distribution of vedo-800CW in affected tissue samples; specifically, the tracer was not located exclusively inside the vessels but also migrated into the gut mucosa. Furthermore, our microscopic analysis confirmed our previous macroscopic findings in which fluorescence intensity was higher in the 15 mg vedo-800CW phase A cohort compared to the therapeutic dose + 15 mg vedo-800CW cohort in phase B. The control group showed only autofluorescent signals (Figure 4A).

**Figure 4:**
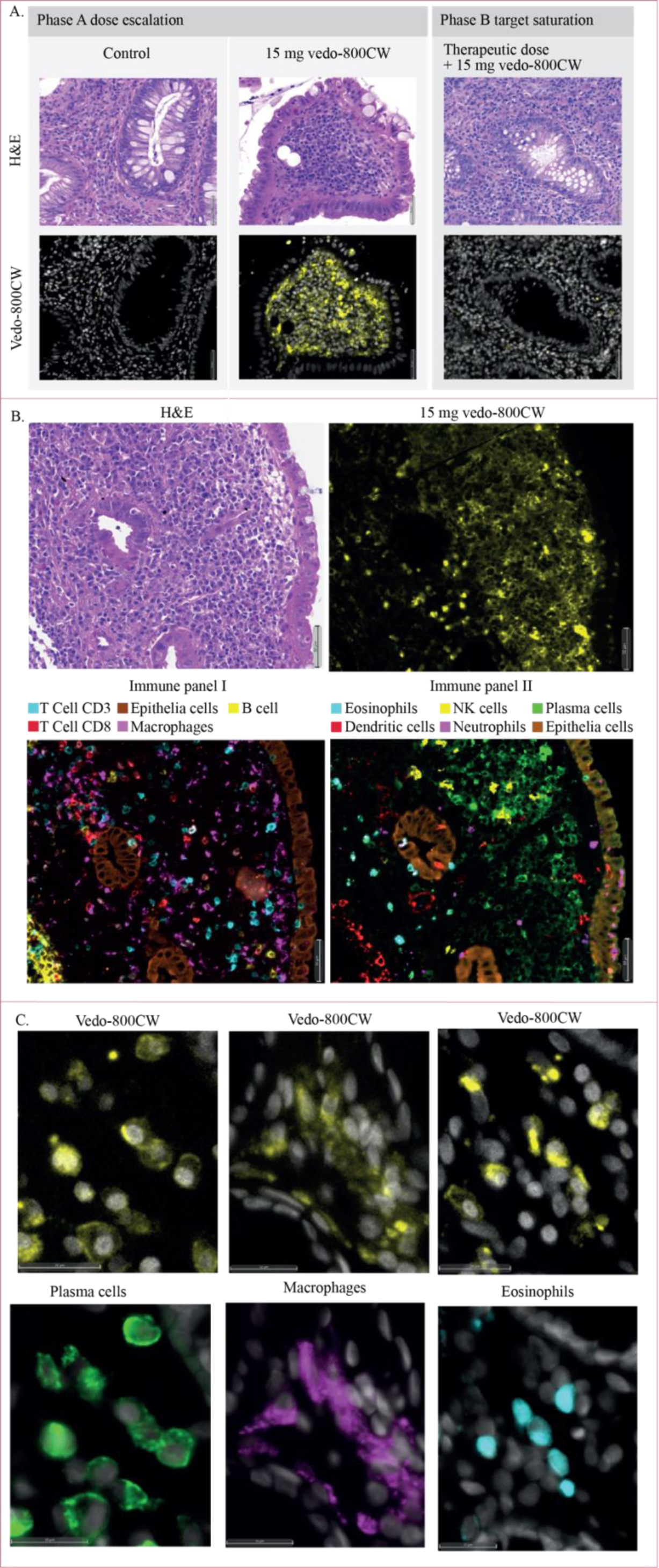
Microscopic distribution of vedo-800CW and target cell identification. (A) Representative fluorescence microscopy images of biopsy sections obtained from patients with active inflammation in the 0 mg cohort (left), the 15 mg cohort (middle), and the therapy + 15 mg cohort (right); the nuclei were counterstained with DAPI shown in gray. Shown above each vedo-800CW fluorescence image is the corresponding H&E-stained section. Note the considerably stronger fluorescence signal in the patient in the 15 mg group compared to the other two patients. (B) Representative images showing an active inflamed H&E section (left) with adjacent the correlation between the distribution of vedolizumab (measured as vedo-800CW fluorescence; right) in tissue sections. The same sections immunostained for two distinct immune panels are shown below. Note that the section stained for immune panel 2 (right) shows an abundance of plasma cells (green) in an area also containing a high number of vedo-800CW‒positive cells (yellow), while the section stained for immune panel 1 (left) shows co-localization between vedo-800CW fluorescence and macrophages (purple). In contrast, there is little correlation between vedolizumab localization and CD3^+^ or CD8^+^ T cells (stained green in the red and blue). (C) Representative high-magnification images showing the spatial correlation between either surface or intracellular vedo-800CW fluorescence (left images in each pair) and specific immune cell types (right images).

To study the drug-target interaction and identify the cell types targeted by vedo-800CW, we performed a more detailed analysis of vedo-800CW binding to a variety of immune cell types ranging from surface binding to internalization of the drug (indicating cytoplasmatic uptake) (Figure 4B). We observed surface binding between vedo-800CW and plasma cells, as well as intracellular localization of vedo-800CW in both eosinophils and macrophages (Figure 4C). Though we did not find clear vedo-800CW positive T cells, CD3+ and CD8+ cells were abundant in regions with strong vedo-800CW signal. Finally, we found no clear evidence of binding between vedo-800CW and any of the other immune cell types studied.

## Discussion

To the best of our knowledge, this is the first study designed to visualize the macroscopic and microscopic distribution of an intravenously administered fluorescent form of vedolizumab in the gut mucosa. Using FMI, we found a dose-dependent increase in vedo-800CW fluorescence in actively inflamed tissues, demonstrating that this approach can be used to assess mucosal drug distribution both *in vivo* and *ex vivo.* We also found that the delivery of unlabeled vedolizumab prior to vedo-800CW administration reduced vedo-800CW fluorescence to the level measured in non-inflamed tissues, indicating specific targeting and suggesting saturation of the target tissue. Furthermore, we found that vedolizumab targets a variety of immune cell types in the inflamed mucosa. Together, these findings represent the first step towards better understanding of the distribution of vedolizumab in the inflamed gut mucosa, thereby helping future research to design studies to investigate the drug’s mechanism of action.

The ability to assess the macroscopic distribution of a drug can be important for confirming that the drug reaches its target site. Moreover, it may also help provide new insights in the drug’s working mechanism and may eventually help predict the patient’s response to therapy. Molecular imaging is considered a powerful tool for increasing our understanding of a drug’s distribution and predicting therapeutic outcome; however, to date these studies primarily involved positron emission tomography (PET) and were performed in an oncology setting.^24,25^ With respect to IBD, Atreya et al. previously reported a study in which they sprayed FITC-labeled adalimumab during endoscopy and then used confocal laser endomicroscopy to identify TNFα-expressing cells.^26^ However, topical application is more of an *in vivo* immunohistochemistry method and does not reflect the drug’s tissue distribution during therapy. Here, we systemically delivered fluorescently labeled vedolizumab by intravenous injection, thus providing visual information regarding its distribution at the site of action, the inflamed mucosa.

Similarly, the ability to quantify local drug levels may be useful in cases in which plasma levels do not reflect the concentration at the site of action. For instance, studies have shown that only ∼5% of a systemically administered drug reaches the inflammatory site or tumor.^27^ In this respect, *in vivo* FMI can be used to qualitatively assess areas with relatively high or low fluorescence intensity, while semi-quantification in biopsies can be used to measure fluorescence intensities *ex vivo*. Moreover, our finding that vedo-800CW remains intact based on SDS-PAGE analysis of fresh-frozen biopsy samples indicates that high fluorescence levels likely correspond to high local drug levels. Our dose-finding study revealed that 15 mg vedo-800CW provided a stronger signal than 4·5 mg and showed a significant difference in fluorescence between actively inflamed tissues and non-inflamed tissues. Interestingly, we also found that binding of vedo-800CW in inflamed tissue was significantly reduced even by a subtherapeutic (75 mg) predose of unlabeled vedolizumab, and this blocking effect was increased further in patients receiving a therapeutic dose of unlabeled vedolizumab. In addition, our *ex vivo* macroscopic fluorescence measurements revealed that the vedo-800CW signal was reduced to the same extent both in patients who received their first therapeutic dose of unlabeled vedolizumab and in patients who previously received more than 14 weeks of therapy. Hence, it is reasonable to speculate that vedolizumab saturates the target tissue even after only a single therapeutic dose. This hypothesis was underscored by our fluorescence microscopy results in patients on vedolizumab treatment, where no specific binding of vedo-800CW to plasma cells was observed. This implies that virtually all α4β7 target molecules were bound by non-fluorescent vedolizumab.

To gain additional insights into a drug’s mechanism of action, it is essential to visualize its microscopic distribution and identify its target cells. With respect to vedolizumab, whether cells other than T cells play a role in its mechanism of action remains an open question.^14,19^ Here, we show that not only is vedo-800CW distributed in a dose-dependent manner in the inflamed mucosa, but it also binds to and/or is taken up by a variety of immune cell types. Specifically, our fluorescence microscopic analysis revealed both surface and intracellular vedo-800CW binding. In addition, consistent with recent findings^14,17,19^ we found that vedolizumab binds to a variety of immune cell types, including plasma cells, macrophages, and eosinophils whereas an actual correlation between T cells and vedo-800CW was not observed. These findings support the hypothesis that T cell migration to the mucosa is prevented and that T cells are likely not the sole therapeutic target of vedolizumab. The potential effect of vedolizumab on plasma cells was described previously. For example, in 1995 Farstad et al. reported an abundance of α4β7-expressing plasma cells in the lamina propria.^28^ Moreover, Canales-Herrerias et al. recently reported in a preprint that vedolizumab can affect the abundance of mucosal plasma cells.^10,19^ Nevertheless, vedolizumab was shown to have opposite effects on the abundance of specific plasma cell subtypes, increasing some subtypes but decreasing others.^10,19^

Here, we found intracellular localization of vedolizumab in both macrophages and eosinophils, consistent with previous studies showing an interaction between vedolizumab and α4β7- expressing macrophages and eosinophils.^12,17,29^ Although this interaction between vedolizumab and various immune cell types is well established, the consequences of this drug binding to different immune cell types remain poorly understood and require further study. Our study has several limitations that warrant discussion. First, due to the relatively low number of vedolizumab-naïve patients and patients enrolled after 14 weeks of vedolizumab therapy, we are unable to draw any conclusions regarding whether FMI can predict the patient’s response to therapy. Ideally, we would have included a group receiving 300 mg vedo-800CW. However, due to production limits of the compound this was not yet feasible. Moreover, we were unable to compare between patients with Crohn’s disease and patients with ulcerative colitis. However, given the complexity of these diseases we believe that combining our approach with more in-depth techniques such as spatial transcriptomics, single cell and RNA sequencing, and gut microbiome analysis may be needed in order to predict response and determine the precise mechanism of action. A second limitation was that our automated quantification of fluorescence intensity during fluorescence microscopy was hampered by the presence of autofluorescence. However, the ability to perform *in vivo* quantification during endoscopy may increase our ability to predict response, and this technique is currently being developed in an ongoing EIC Horizon Pathfinder project (grant number 101046923).

Our approach using FMI to visualize fluorescently labeled drugs in inflammatory diseases can also be used to gain insights into drug distribution and identify target cells in other contexts. For example, our group is currently using FMI to visualize fluorescently labeled adalimumab and ustekinumab in arthritis, psoriasis, and IBD (ClinicalTrials.gov trials NCT03938701 and NCT05725876). Using a similar approach may also be valuable in early drug development trials. Measuring the local drug concentration, identifying the drug’s target cells, and determining target saturation can provide important information for making go/no-go decisions during development.^30^ Thereby, accelerating the drug development process, decreasing costs, and allowing for more accurate patient selection when new drugs enter the market.

In conclusion, this phase I feasibility study using a novel optical imaging approach provides the first detailed information regarding the macroscopic and microscopic distribution of vedolizumab in the inflamed gut, including information regarding its target cells. Our stepwise FMI approach including *in vivo* fluorescence endoscopy, *ex vivo* fluorescence analysis of biopsies, and fluorescence microscopy has the potential to increase the understanding of the underlying mechanism of action, the drug’s distribution, and its targets, leading to the optimization of treatment in individual patients.

## Abbreviations

a.u.: arbitrary units
GMP: good manufacturing practice
CD: Crohn’s disease
FFPE: formalin-fixed, paraffin-embedded
FI: fluorescence intensity
FMI: fluorescence molecular imaging
H&E: hematoxylin and eosin
IHC: immunohistochemistry
UC: ulcerative colitis
UMCG: University Medical Center Groningen.

## Acknowledgments

This work has received funding from the Innovative Medicines Initiative 2 Joint Undertaking (JU) under grant agreement No 831514 (Immune-Image). The JU receives support from the European Union’s Horizon 2020 research and innovation programme and EFPIA. These funding sources played no role in the study design, data collection, data analysis, decision to publish, or preparation of the manuscript. The authors would like to thank lab technicians Gert Jan Meersma and Manon Buist-Homan for performing all *ex vivo* staining procedures. In addition, the authors would like to thank English Editing Solutions and Sieben Medical Art for editing the manuscript. The authors would also like to thank all of the patients who participated in this study.

## Author contributions

*Guarantor of the article*: Wouter B. Nagengast

*Specific author contributions*: All authors were involved in conceptualization and study design. WBN was responsible for funding acquisition and resources. RYG, AMvdW, EAMF, and GD were responsible for patient enrollment. RYG, MDL, AMvdW, and PV performed all *in vivo* study procedures. RYG, MDL, AMvdW, and PV performed all *ex vivo* imaging procedures. MDL was responsible for tracer production, quality control, and release. GK-U analyzed the H&E-stained sections for inflammation criteria (blinded with respect to the fluorescence result). RYG, AMvdW, and PV contributed to the interpretation and analysis of the imaging results. MD and SJ performed microscopic drug imaging and histological tissue analysis at the Regeneron Clinical Histology Core. RYG, MDL, AMvdW, and PV wrote the first draft of the manuscript with input from all other authors. MD, SJ, MAH, MNLdH, EAMF, GD, GK-U, and WBN interpreted the results and critically reviewed the manuscript. All authors approved the final version of the manuscript, including the authorship list.

## Data sharing

The main data supporting the outcomes of this clinical trial are available within the manuscript and its supplementary materials. The study protocol and de-identified raw FMI data may be obtained by sending an e-mail to the corresponding author including a clear research proposal. Data requestors will need to sign a data access agreement in order to gain access.

