## Supplementary material for "Using fluorescently labeled vedolizumab to visualize local drug distribution during colonoscopy and identify mucosal target cells in patients with inflammatory bowel disease": Table S1

**Table S1: Overview of the markers, antibodies used for immunofluorescence, and target cells in the two immune panels.**

| IHC assay | Marker | Ab clone | Fluorophore | Vendor | Aimed target cells |
|---|---|---|---|---|---|
| Panel 1 | CD20 | SP32 | Rhodamine 6G | Abcam | B cells |
| | CD3 | SP162 | DCC | Abcam | CD3+ T cells |
| | CD8 | SP239 | RED 610 | Abcam | CD8+ T cells |
| | CD68 | SP251 | CY5 | Abcam | Macrophages |
| | Foxp3 | SP97 | FAM | Abcam | CD3+CD8-FOXP3+ |
| Panel 2 | BCMA | E6D7B | Rhodamine 6G | Cell Signal Technology | Plasma cells |
| | ECP | EPR20357 | DCC | Abcam | Eosinophils |
| | PanCK | AE1/AE3/PCK26 | RED 610 | Ventana (Roche) | Epithelial cells |
| | MPO | EPR20257 | CY5 | Abcam | Neutrophils |
| | CD11c | EP1347Y | FAM | Abcam | Dendritic cells |
