## Supplementary material for "Using fluorescently labeled vedolizumab to visualize local drug distribution during colonoscopy and identify mucosal target cells in patients with inflammatory bowel disease": Figure S1

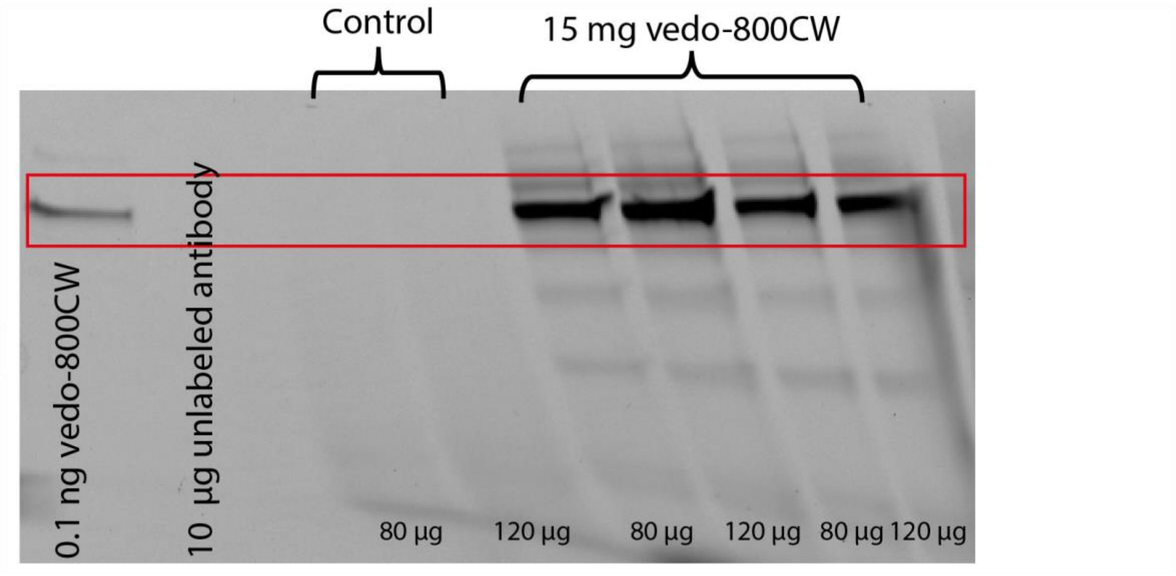


**Figure S1: Vedolizumab-800CW integrity**

The SDS-PAGE experiment performed on fresh-frozen patient biopsy samples determined the stability and integrity of the vedo-800CW conjugate.
