## Supplementary material for "Using fluorescently labeled vedolizumab to visualize local drug distribution during colonoscopy and identify mucosal target cells in patients with inflammatory bowel disease": Figure S2

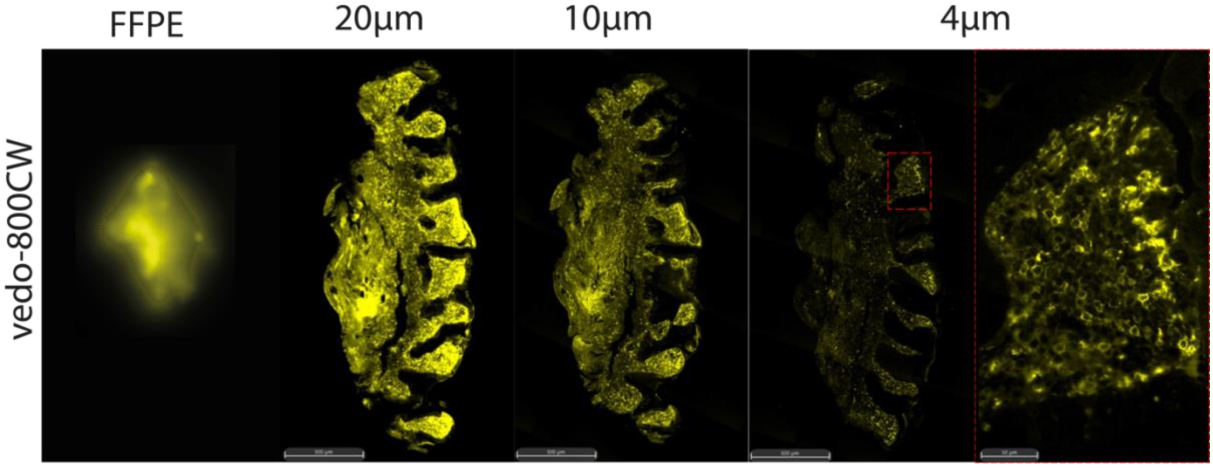


**Figure S2: From macroscopic to microscopic ex vivo imaging**

Images of different biopsy thicknesses from a patient receiving 15 mg vedo-800CW. From left to right different thicknesses are shown; from the whole formalin fixed paraffin embedded (FFPE) biopsy to the 4 µm tissue section revealing the intensity of the vedo-800CW fluorescence signal. The FFPE section is scanned with the Odyssey CLx flatbed scanner (LI-COR Biosciences). Further images were generated using a Zeiss AxioScan Z1 slide scanner.
